# ADHERENCE TO NATIONAL HEALTHCARE REFERRAL GUIDELINES AND ITS EFFECT ON THE MANAGEMENT OUTCOMES AMONG CHILDREN SEEN AT A TEACHING HOSPITAL IN WESTERN KENYA

**DOI:** 10.1101/2020.11.11.20229575

**Authors:** Paul Jairus Njanwe, Irene Marete, Samuel Ayaya

**Affiliations:** Department of Child Health and Paediatrics, School of Medicine – Moi University, Kenya

**Keywords:** Hospital Referral, Tertiary Referral Center, Self-Referral, Physician, Paediatrics

## Abstract

**Introduction:** Referral guidelines are meant to ensure coordination and continuity across all levels of healthcare. Poor adherence to these guidelines could result in increased morbidity and mortality among the patients who are denied access; especially in the resource constrained healthcare settings in developing economies.

**Aim:** To determine adherence to the national healthcare referral guidelines and immediate outcomes of children seen at a tertiary teaching hospital in Western Kenya.

**Materials and methods:** A Cross-sectional study conducted at the Pediatric emergency department of Moi Teaching and Referral Hospital in Western Kenya between February to June 2016. A total of 422 children aged below 15 years were recruited systematically. Sociodemographic and clinical data were collected using interviewer administered questionnaires and clinical chart reviews respectively. Checklists were used to collect information from ambulances. Pearson chi-square tests and odds ratios were used to test for association between predictor and outcome variables using statistical package for social science (SPSS) version-24.

**Results:** More than half (55.5%) of the 422 children enrolled were male while 51.4% were aged between 5 to 14 years. Hospital referrals accounted for 15.9% (n=67) with the rest being self-referrals and no counter referrals seen. Adherence to all the four transfer guideline requirements was observed in 46.3% (n=31) of the 67 hospital referrals. Less than half (46.3%) of the hospital referrals had their referring facilities calling the receiving facility prior to initiating the referral; 83.6% had a referral document; 64.2% were transferred in ambulances while 68.7% (n=46) were accompanied by health care workers. Most (88.1%) of the hospital referrals were admitted. Lower level of parental education (p= 0.025), residing outside the host county (p<0.001) and a child being older than five years (p = 0.015) were significantly associated with hospital referrals. Hospital referrals were nearly three times (AOR = 2.932; 95% CI: 2.422 – 3.550; p<0.001) more likely to be admitted compared to children who were self-referred.

**Conclusion:** There is low adherence to national healthcare referral guidelines among children seen at the second largest national hospital in Kenya; with less than half of hospital referrals transferred as per the transfer process guidelines.

## Introduction

The process of transferring patients from lower-level healthcare systems to those with advanced infrastructural and human resources is a key element of the health care system globally. In low- and middle-income countries such as Kenya, this is necessitated by lack of adequate technological and infrastructural resources as well as professional skills to handle some debilitating health conditions. Patient referrals especially between various hospitals by healthcare providers acts as a building elevator to facilitate forward and backward management of clients’ needs. A functional patient referral system ensures optimal use of healthcare facilities and personnel by improving communication among all healthcare providers involved in the patient’s management [1]. The healthcare system in Kenya is hierarchical from the community health care services, primary health care, county referral services to the national teaching and referral facilities[2]. The higher the level of the facility, the more sophisticated it is in providing diagnostic, therapeutic and rehabilitative services (Tom Kizito et al, 2005).

To ensure proper coordination of the patient referral system, Kenya’s Ministry of Health developed National Healthcare Referral Guidelines [3]. Both patients and healthcare providers are expected to adhere to these referral guidelines and ensure safe and proper patient transfer across healthcare facilities. Optimal adherence to these guidelines further ensures that patients receive the full spectrum of care provided by the health system, regardless of the level at which they physically access health care. The referral guidelines recommend that patients should first seek medical care at primary healthcare facilities except in emergency situations where referral facilities can be accessed directly. In the event of hospital to hospital referral, the attending healthcare provider is required to call the receiving facility in advance to ensure availability of the required medical service, legibly fill-out a client referral form in English, indicate previous medical care offered including attaching all the relevant diagnostic test results, especially in emergency referrals. Adherence to transfer process guidelines especially among pediatric patients requires that the patient is transferred in an ambulance with a functional oxygen supply, first-aid kit, essential medicines and a transfer couch. The patient in the ambulance must also be accompanied by a competent healthcare provider.

Despite all these guidelines in place, adherence to referral and transfer process guidelines by both patients and healthcare providers has been a challenge in many national referral hospitals. This has been exemplified by long queues of patients with medical conditions that could be easily managed at primary healthcare facilities. Parents of paediatric patients living in urban centers near these national referral hospitals often opt to bypass the healthcare hierarchy by self-referring their children.

This study therefore aimed to determine the adherence to National Healthcare Referral Guidelines and document immediate outcomes among children seen at Moi Teaching and Referral Hospital (MTRH) in Eldoret-Kenya. Specifically, it determined the proportion of children seen at MTRH who were referred from other health facilities; described the patterns of referrals; determined the level of adherence to the transfer process guidelines and the immediate management outcomes of children seen.

## Materials and methods

This was a cross-sectional study conducted among paediatric patients attending MTRH’s sick child clinic February to June 2016. The hospital is the second largest national government teaching and referral hospital in Kenya with a bed capacity for 800 patients and attends to patients in the greater Western Kenya. The study design was adopted because the researcher only came into contact with the children and their parents at enrollment, while all other information were obtained from medical records. The eligible children were those below 15 years who were not revisiting the sick child clinic for review or follow-up due to the same condition during the study period. Referred children who died on arrival at the sick-child clinic were excluded. The children were further sampled and enrolled systematically with a sampling interval of 28 until the desired sample size of 422 was achieved.

Patient data was collected using an interviewer administered questionnaire that was divided into six sections: demographic data, referral status, referral process, status of the transferring vehicle or ambulance, referral documents, and care given at MTRH. If the patient was self-referred, information such as the nearest health facility, and distance from MTRH was collected. Primary data source was the parents or guardians of the sick children; while secondary data obtained from referral notes, referral forms, patient transfer forms and medical charts. All the patients received standard pediatric care as is required by the ethical guidelines and approvals obtained from the Institutional Research and Ethics Committee (IREC) of MTRH and Moi University School of Medicine (Approval # 1516). Other ethical considerations such as parental consent and paediatric assents were obtained prior to data collection as well as participants privacy and confidentiality was ensured by deidentifying patient data and storing their information in password protected databases. Descriptive (frequency, mean and median with corresponding proportions, standard deviations and interquartile ranges) and Inferential (Pearson chi-square and odds ratios at 95% confidence interval) statistical analysis were conducted using Statistical Package for Social Sciences (SPSS) version 24.

## Results

This study enrolled 422 children with more than half (51.4%; n=217) of them older than 5 years of age. The male to female ratio was 1.2:1 and majority (88.9%; n=375) of the children lived in the hospital’s host county of Uasin Gishu in Western Kenya (Table/Fig 1).

More than one-tenth (15.9%; n=67) of the children enrolled were hospital referrals to MTRH, while the rest were self-referrals with no counter-referrals reported. Among the hospital referrals, majority (86.6%; n=58) were from government facilities, followed by those from private hospitals (11.9%; n=8), with the least representation from private clinics at 1.5% (n=1). The main reason for referral was to seek specialized care.

The main referral patterns studies were the proportion of hospital referrals living near a public hospital, the distance they covered to reach MTRH, their chief complaints, tests done prior to referral (Table/Fig. 2). When a test of association was conducted, it was determined that children whose parents or guardians had secondary education or less, lived outside Uasin Gishu county were more than five (5) years of age were more likely to be referred from healthcare facilities (Table/Fig. 3).

When adherence to transfer guidelines was assessed, four aspects (Calling prior to referral, having a Referral Document, being transferred by an ambulance and the patient being accompanied by a healthcare worker) were scored. Lack of adherence to any of the steps was scored as zero while partial adherence was defined as compliance to one of the four transfer guidelines. Total adherence was when the child’s transfer adhered to all the four aspects. When this technique was adopted, 14.9% (10) of the children referred were transferred without adhering to any of the four transfer guidelines, while nearly half (46.3%; n=31) of all hospital referrals had total adherence. More than four-fifths of the children referred from other health facilities came to MTRH with a referral document, all the ambulances had oxygen supply that was intact (Table/Fig. 4).

Majority of the hospital referrals were admitted while nearly two-thirds of the self-referrals were treated and discharged (Table/Fig. 5). Paediatric patients who were referred from hospitals were nearly three times (AOR = 2.932; 95% CI: 2.422, 3.550) more likely to be admitted to the wards than those who were not (Table/Fig. 6).

## Discussion

### Proportion of Hospital Referrals

Previous studies have demonstrated that low proportions of children are often referred to tertiary national and teaching hospitals from lower-level medical facilities [4–6]. In this study, 15.9% of the children seen at MTRH were hospital referrals as required by the national healthcare referral guidelines. This reported proportion of hospital referrals in Kenya among paediatric patients is less than a third of that reported in Canada at 45.5% [7]. Studies have also compared the proportion of referrals in the United States of America (USA) and the United Kingdom (UK) among mixed populations of children and adults (0-64 years) with varying proportions. In the USA, 30% to 36.8% of patients were referred from medical facilities [8] compared to only 13.9% in the UK [9]. This difference is attributed to the lower proportion of paediatric specialists in the UK in comparison to the United States of America [8,10].

The findings of this study differ from other published studies conducted in East Africa. In a study conducted at Tanzania’s Kilombero District Health Care System [11]; out of 5,030 new pediatric cases from government and second level health facilities, 28 (0.6%) were referred for specialized care. This very low proportion of referrals in Tanzania was attributed to the fact that accurately ill children are not often brought to the health facilities; health facility staff do not identify children who need referral and healthcare workers only refer children with socioeconomic support to travel to the referral health facility. Due to similarity in the socioeconomic status of the parents to paediatric patients from both Kenya and Tanzania, this could explain the similarity in low proportions of hospital referrals for paediatric patients in Western Kenya.

### Patterns of referrals

This study assessed living near a public hospital, distance to the referral facility, the chief complaint of the paediatric patient, tests done prior to referral, reasons given for referral and the final diagnosis as patterns of paediatric patient referral. Nearly two-thirds (62%) of those referred lived near a public hospital which was four times higher than 15.7% reported in South Africa [12]. Majority of the children referred from other health facilities had to travel more than 10 kilometers to seek care. Living far away from the national referral hospital increased the likelihood of the child being referred from a different medical facility compared to those living near the referral hospital who were self-referred.

The most common symptom in nearly half (47.8%) of the paediatric patients referred from other health facilities presented with a fever, which differs from the 17.2% of those seen in a Hong Kong referral hospital who were wheezing [13]. This difference could be attributed to socioeconomic and environmental differences in Kenya and Hong Kong. Whereas Kenya has more cases of infections reported in children and presenting as fever[14], wheezing could be common in Hong Kong due to a high prevalence of asthma.

Most (79.1%) of the children were referred to MTRH for specialized care while in Hong Kong most of the children were referred due to growth problems. This is because in Kenya, there is an insufficiency of medical infrastructure and specialists in many public hospitals in the counties [3]. This necessitates referral to the national hospitals such as MTRH for specialized care. On arrival at the receiving referral hospital, the children were diagnosed with anemia (15.6%) and pneumonia (10.4%). This proportion of pneumonia diagnosis reported in this study was lower than that reported at an advanced paediatric emergency care in Vietnam [4], where 23.7% of the children were diagnosed with pneumonia.

### Adherence to transfer process guidelines

This study reports that nearly half (46.3%; n=31) of the children referred were transferred in total adherence to all the steps in the transfer guidelines. The steps of interest were: the referring facility was required to call the receiving facility prior to referral; having a referral document (either a referral form or a referral note);referral by an ambulance (that has essential medicines, first aid kit, transfer couch and an oxygen supply that is both intact and functional); and the patient should be accompanied (by either a clinical officer or a registered nurse). Majority of the children (83%) either had a referral form or referral note as a referral document. This contrasts findings from Saudi Arabia [15] where all the children referred from primary care to hospitals had a referral document. This disparity could be attributed to poor communication channels between the referring and receiving facilities in Kenya. In Punjab-India [16], pre-referral documentation was also found to be inadequate and lower than those reported here at 3.7%. The proportions of children who were transferred by an ambulance (64.2%) and those who were accompanied (68.7%) was close to those reported in Vietnam [4] at 57.8% and 49.6% respectively. Higher government ambulance transfer rate of 85.5% from public hospitals was reported in India [16]. Furthermore, in this study, 87% of the children transferred in an ambulance were accompanied by a nurse compared to 25.1% in Vietnam [4]. Although no child in the current study was accompanied by a medical officer, 7.6% of those in Vietnam were [4]. This difference could be attributed to the variance in the proportional distribution of human resources for health in the two countries.

### Management outcomes

More than one third (39.1%) of this study’s participants who were hospital referrals ended up being admitted. This finding was consistent with that of Habib et al (2017) which reported admission outcomes at 39.3%.However, this was four-times higher than that in Vietnam [4]. A low proportion (0.5%) of the referred children in this study died comparable to that in Afghanistan at 3% [17]. Furthermore, nearly all (88%) of the children referred from facilities were admitted in contrast with a study in Saudi Arabia [15] where less than half of the referrals were treated and discharged. This may be because most of the referred patient at MTRH sick child clinic were sicker and more required emergency inpatient management.

## Conclusions and Recommendations

This study determined that less than a quarter of paediatric patients at a national referral hospital in Western Kenya were hospital referrals with the majority being self-referrals. There was less than 50% adherence to transfer process guidelines among hospital referrals. Nearly all the hospital referrals were admitted for further management with approximately three-time likelihood of admission compared to self-referred children who were mostly treated and discharged. There is need for reduction in the proportion of self-referrals to national referral hospitals through targeted improvement on adherence to national healthcare referral guidelines. Most of the children seeking care at the national referral hospital should be encouraged to visit primary and secondary level healthcare facilities to reduce on self-referrals. Referring health-facilities should be encouraged to totally comply with the recommended transfer processes. Because this study did not determine the perceptions of the parents or guardians and reasons for self-referring their children; further qualitative studies determining reasons for self-referrals and lack of adherence to transfer guidelines should be conducted.

## Data Availability

All the data obtained from this study will be made publicly available

## APPENDICES

**Table/Fig. 1:** Sociodemographic characteristics of Study Participants.

| Variable | Category | Frequency | Percent |
| --- | --- | --- | --- |
| Gender of the child | Male | 234 | 55.5 |
|  | Female | 188 | 44.5 |
| Age of the child | <1year | 51 | 12.1 |
|  | 1 – 5years | 154 | 36.5 |
|  | 6 -14years | 217 | 51.4 |
| Occupation of parent/guardian | Formally Employed | 105 | 24.9 |
|  | Informally Employed | 199 | 47.2 |
|  | Unemployed | 118 | 27.9 |
| Monthly Family/household Income (in KSh) | <10000 | 126 | 29.9 |
|  | 10000 – 20000 | 211 | 50.0 |
|  | 21000 – 50000 | 71 | 16.8 |
|  | 51000 - 100000 | 12 | 2.8 |
|  | >100000 | 2 | 0.5 |
| Residence | Uasin Gishu county | 375 | 88.9 |
|  | Other counties | 47 | 11.1 |
| Parent/Guardian level of Education | None | 4 | 0.9 |
|  | Primary | 65 | 15.4 |
|  | Secondary | 274 | 64.9 |
|  | Tertiary | 79 | 18.7 |

**Table/Fig. 2:**
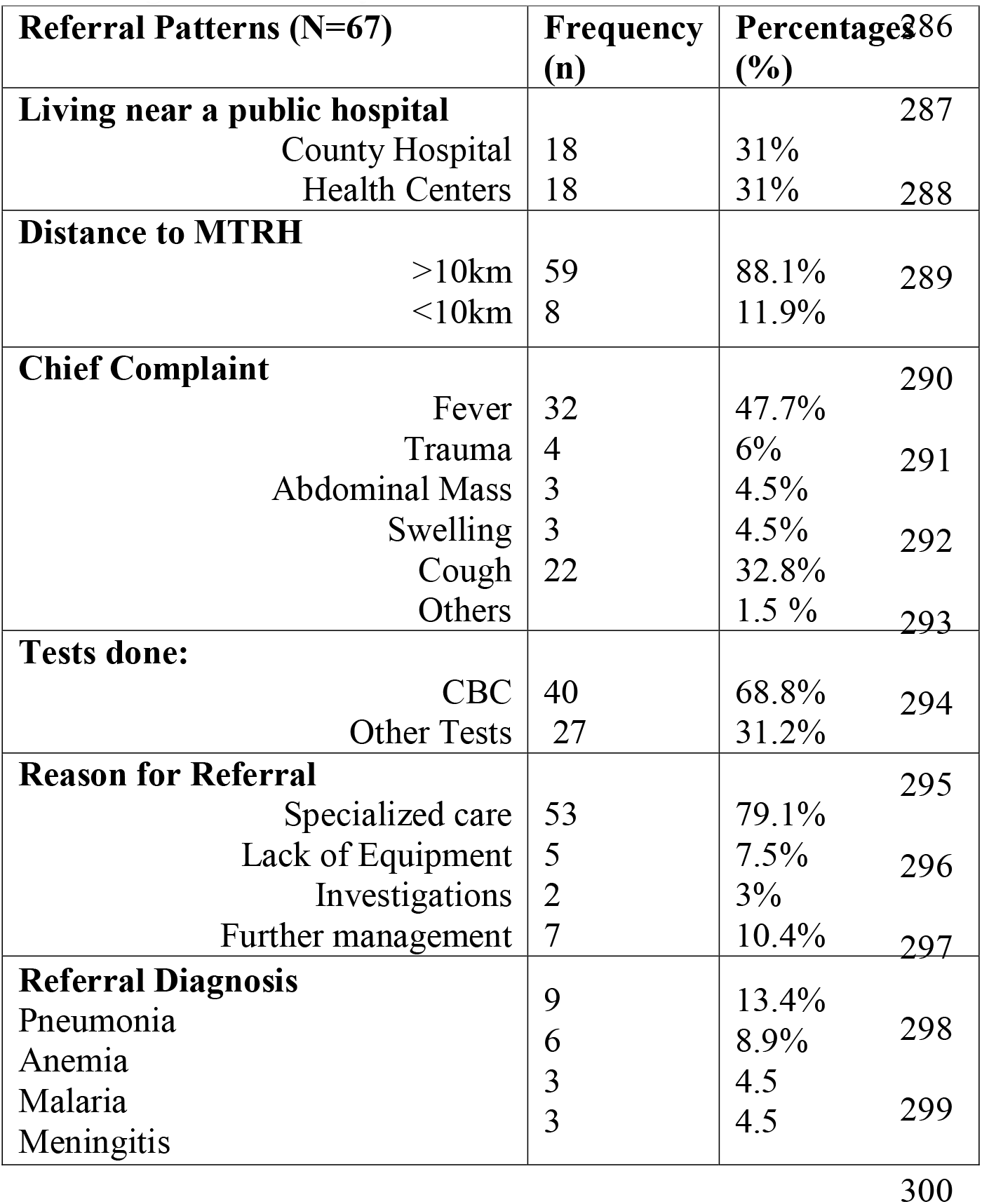
Summary of Patient Characteristics of Children Referred to MTRH.

| Referral Patterns (N=67) | Frequency (n) | Percentage (%) |  |
| --- | --- | --- | --- |
| **Living near a public hospital** |  |  | 287 |
| County Hospital | 18 | 31% |  |
| Health Centers | 18 | 31% | 288 |
| **Distance to MTRH** |  |  |  |
| >10km | 59 | 88.1% | 289 |
| <10km | 8 | 11.9% |  |
| **Chief Complaint** |  |  | 290 |
| Fever | 32 | 47.7% |  |
| Trauma | 4 | 6% | 291 |
| Abdominal Mass | 3 | 4.5% |  |
| Swelling | 3 | 4.5% | 292 |
| Cough | 22 | 32.8% |  |
| Others |  | 1.5 % | 293 |
| **Tests done:** |  |  |  |
| CBC | 40 | 68.8% | 294 |
| Other Tests | 27 | 31.2% |  |
| **Reason for Referral** |  |  | 295 |
| Specialized care | 53 | 79.1% |  |
| Lack of Equipment | 5 | 7.5% | 296 |
| Investigations | 2 | 3% |  |
| Further management | 7 | 10.4% | 297 |
| **Referral Diagnosis** |  |  |  |
| Pneumonia | 9 | 13.4% |  |
| Anemia | 6 | 8.9% | 298 |
| Malaria | 3 | 4.5 |  |
| Meningitis | 3 | 4.5 | 299 |

**Table/Fig. 3:** Association between Sociodemographic Characteristics and Facility Referral.

| Characteristic | AOR (95% CI) | p-value |
| --- | --- | --- |
| **Level of Education (Parent/Guardian):** |  |  |
| ≤ Secondary | **1.146 (1.046 – 1.256)** | **0.025** |
| Tertiary | 0.435 (0.198 – 0.960) |  |
| **County of Residence** |  |  |
| Uasin Gishu | 0.461 (0.353 – 0.601) | **<0.001** |
| Other Counties | **19.604 (10.256 -37.476)** |  |
| **Pediatric Age Group** |  |  |
| ≤ 5 years | 0.713 (0.526 – 0.968) | **0.015** |
| > 5 years | **1.372 (1.091 – 1.727)** |  |

**Table/ Fig. 4:** Adherence to Transfer Guidelines among Facility Referrals (N=67)

| REFERRAL GUIDELINE | YES<br>n(%) | NO<br>n(%) |
| --- | --- | --- |
| Calling prior to Referral | 32 (47.8%) | 35 (52.2%) |
| Referral Document | 56 (83.6%) | 11 (16.4%) |
| *Referral form:* | 31 (55.4%) |  |
| *Referral note:* | 25 (44.6%) |  |
| Transfer by an Ambulance | 43 (64.2%) | 24 (35.8%) |
| *Oxygen supply:* | 43 (100%) |  |
| *Oxygen supply intact:* | 43 (100%) |  |
| *Equipment functional:* | 40 (93%) | 3 (7%) |
| *Essential Medicines:* | 38 (88.4%) | 5 (11.6%) |
| *First Aid Kit:* | 40 (93%) | 3 (7%) |
| *Transfer Couch:* | 42 (97.7%) | 1 (2.3%) |
| Patient Accompanied | 46 (68.7%) | 21 (31.3%) |
| *Clinical officer:* | 6 (13%) |  |
| *Enrolled Community Nursing:* | 11 (23.9%) |  |
| *Registered Nurse:* | 28 (60.9%) |  |
| *Nursing student:* | 1 (2.2%) |  |

**Table/Fig. 5:** Management outcomes of pediatric patients referred to MTRH (N=422)

|  | Referral Pattern |  | Total |
| --- | --- | --- | --- |
|  | Self-Referral | Hospital-Referral |  |
| Admitted to The Ward | 106 (29.9%) | 59 (88.1%) | 165 (39.1%) |
| Death | 1 (0.3%) | 1 (1.5%) | 2 (0.5%) |
| Referred to Specialized Clinic | 25 (7%) | 2 (3%) | 27 (6.4%) |
| Discharged Home | 223 (62.8%) | 5 (7.4%) | 228 (54%) |
| Total | 355 (100%) | 67 (100%) | 422 (100%) |

**Table/Fig. 6:**
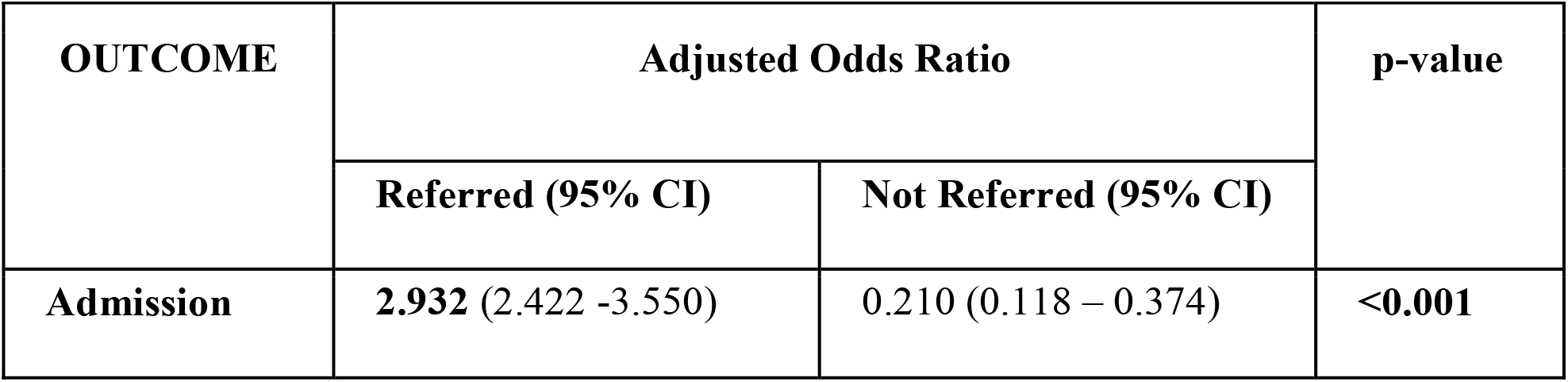
Association between Referral Status and Admission.

| OUTCOME | Adjusted Odds Ratio |  | p-value |
| --- | --- | --- | --- |
|  | Referred (95% CI) | Not Referred (95% CI) |  |
| Admission | 2.932 (2.422 -3.550) | 0.210 (0.118 – 0.374) | <0.001 |

